# The competing risk between in-hospital mortality and recovery: A pitfall in COVID-19 survival analysis research

**DOI:** 10.1101/2020.07.11.20151472

**Authors:** Abderrahim Oulhaj, Luai A. Ahmed, Juergen Prattes, Abubaker Suliman, Ahmed R. Alsuwaidi, Rami H. Al-Rifai, Harald Sourij, Ingrid Van Keilegom

**Affiliations:** Institute of Public Health, College of Medicine and Health Sciences, United Arab Emirates University, Al Ain, United Arab Emirates; Zayed Center for Health Sciences, United Arab Emirates University, United Arab Emirates; Department of Internal Medicine, Section of Infectious Diseases and Tropical Medicine,Medical University of Graz, Austria; Department of Pediatrics, College of Medicine and Health Sciences, United Arab Emirates University, Al Ain, United Arab Emirates; Department of Internal Medicine, Division of Endocrinology and Diabetology, Medical University of Graz, Austria; ORSTAT, KU Leuven, Belgium

**Keywords:** COVID-19, competing risk, in-hospital mortality, survival analysis, recovery

## Abstract

**Background:** A plethora of studies on COVID-19 investigating mortality and recovery have used the Cox Proportional Hazards (Cox PH) model without taking into account the presence of competing risks. We investigate, through extensive simulations, the bias in estimating the hazard ratio (HR) and the absolute risk reduction (ARR) of death when competing risks are ignored, and suggest an alternative method.

**Methods:** We simulated a fictive clinical trial on COVID-19 mimicking studies investigating interventions such as Hydroxychloroquine, Remdesivir, or convalescent plasma. The outcome is time from randomization until death. Six scenarios for the effect of treatment on death and recovery were considered. The HR and the 28-day ARR of death were estimated using the Cox PH and the Fine and Gray (FG) models. Estimates were then compared with the true values, and the magnitude of misestimation was quantified.

**Results:** The Cox PH model misestimated the true HR and the 28-day ARR of death in the majority of scenarios. The magnitude of misestimation increased when recovery was faster and/or chance of recovery was higher. In some scenarios, this model has shown harmful treatment effect when it was beneficial. Estimates obtained from FG model were all consistent and showed no misestimation or changes in direction.

**Conclusion:** There is a substantial risk of misleading results in COVID-19 research if recovery and death due to COVID-19 are not considered as competing risk events. We strongly recommend the use of a competing risk approach to re-analyze relevant published data that have used the Cox PH model.

## 1. Introduction

Severe acute respiratory syndrome coronavirus 2 (SARS-CoV-2), the cause of the pandemic Coronavirus Disease 2019 (COVID-19), has affected more than 13.0 million people in over 190 countries and caused more than 570,000 deaths, as of July 13, 2020^1^. Numerous retrospective or prospective cohort studies and randomized clinical trials (RCTs) were conducted to evaluate different therapeutic interventions^2-7^. The results of these studies have not only impacted treatment strategies in COVID-19 patients, but have also influenced health-policy decision making including continuation, modification, or termination of the use of some of the studied drugs including Hydroxychloroquine, Remdesivir or convalescent plasma therapy^3 5 7^.

The statistical methods commonly used in these studies to investigate primary outcomes such as in-hospital mortality or recovery are based on the standard Cox proportional hazards model (Cox PH) and the Kaplan–Meier estimator^8 9^. When in-hospital mortality due to COVID-19 is the outcome of interest, these two methods implicitly treat recovered patients as right-censored. Similarly, when recovery from COVID-19 is the outcome of interest, these two methods consider patients who died as right-censored. This way of proceeding implies that patients who recovered (respectively died) have similar risk of death (respectively recovery) compared to those still at risk (i.e. still hospitalised). In fact, death due to COVID-19 and recovery are mutually exclusive competing events, and therefore, recovery (respectively death) should be considered as a competing risk for death (respectively recovery) rather than right-censored. A competing risk is an event whose occurrence precludes the occurrence of the event of interest^10^. Ignoring the competing risk will usually lead to estimates of hazard ratios (HR) and absolute risks that are largely biased^11 12^ and eventually misleading conclusions. If we would be interested in overall death from any disease (as opposed to death due to COVID-19), then a competing risk approach would not be necessary since recovery is in that case not a competing event but is rather censoring the event of interest.

Despite the abundance of statistical papers recommending the use of survival models that account for competing risks^11 13^, these models are still underused in many medical studies especially in COVID-19 related research. The main objective of this study is to investigate, through extensive simulations, the bias that occurs when estimating the HR and the absolute risk reduction (ARR) using the Cox PH model in the presence of competing risks. In this simulations, the Fine and Gray (FG) model^14^, which takes competing risks into account, is used as an alternative to the Cox PH model.

## 2. Methods

We simulated data for a fictive clinical trial in COVID-19 patients where a given treatment is compared to placebo. In this simulation, 10,000 patients were randomly assigned to receive either the treatment or a placebo in a 1:1 ratio. We have chosen such a big sample size in order to discard any justification related to small sample sizes. In this simulated data, patients are entered in the study at the date of randomization and are followed-up for a maximum length of stay in the hospital up to 50 days. During that period, patients can either die, recover from COVID-19, withdraw from the study, are lost to follow-up or reach the end of study with no event.

The primary outcome is the time from randomization until death due to COVID-19. Recovery during the follow up period is considered as a competing event, while time to loss to follow-up, withdrawal or reaching the end of the study with no event are considered as right censoring. Note that in doing so, we ignore cases where a patient dies from COVID-19 after recovery. This, if it happens, is in fact very rare and would not affect the current results.

The main quantities of interest to be estimated and investigated are a) the hazard ratio (HR) of the primary outcome defined as the hazard of death due to COVID-19 in treated patients divided by the hazard of death in the placebo group, and b) the 28-day ARR of in-hospital mortality defined as the risk of death due to COVID-19 within 28 days in the treated group minus the risk of death within 28 days in the placebo group.

### Data generating process

We simulated the data according to six scenarios as described in table 1. The scenarios represent different situations of the treatment effect on the primary outcome (COVID-19 related death) and its competing event (Recovery). Scenario 6, for instance, represents the situation where the treatment has an effect on both death and recovery but the effect on recovery is higher than the one on death. For each scenario, 1,000 samples each of size 10,000 patients (5,000 treated and 5,000 placebo) were generated from different data generating processes (DGP). More specifically, the times to death in each sample were generated from a proportional hazards model with baseline hazard coming from a mixture of a point mass at infinity and an exponential variable truncated at 50 days, and the finite recovery times were generated from another proportional hazards model, again truncated at 50 days. Time to right-censoring were generated from a mixture of a uniform distribution and a point mass distribution at 50 days. This distribution for right-censoring was chosen in a way that fewer patients withdraw during the first days of the trial and others to remain alive by the end of the study.

**Table 1:**
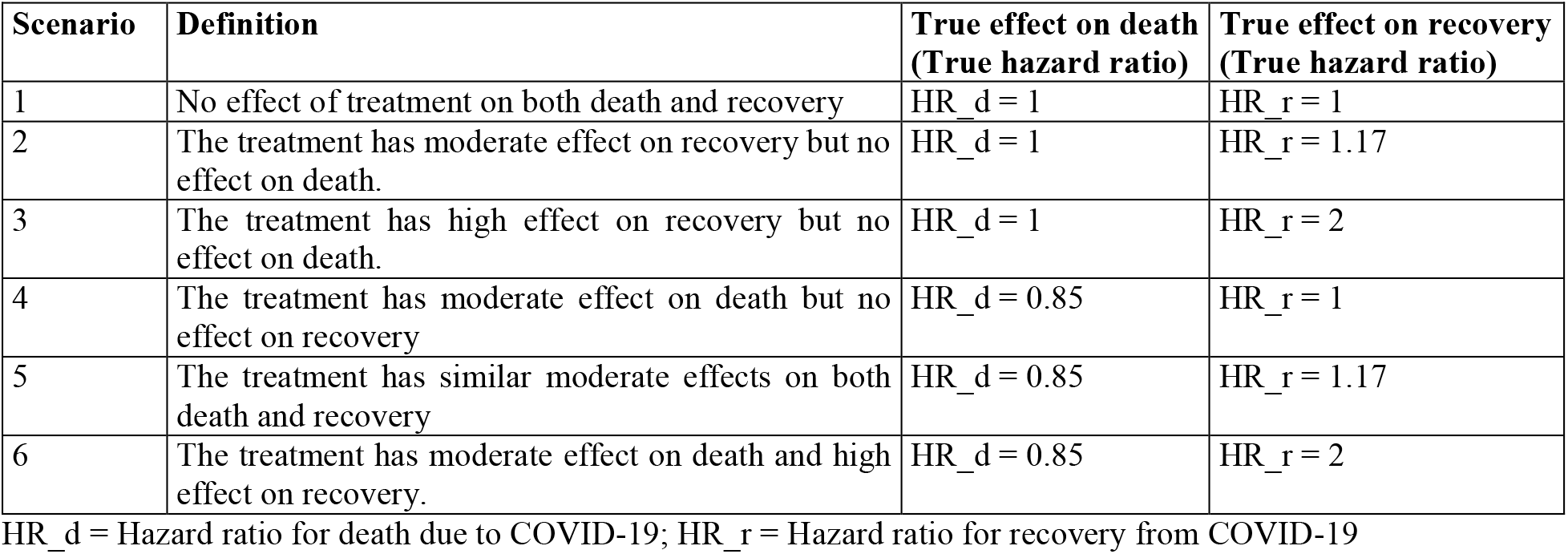
The six scenarios used in the simulations.

We also allowed the chance of recovery and the median time to recovery within each scenario to vary leading to different DGPs. More specifically, the chance of recovery in the placebo group was set to be 5%, 80%, 90% and 95%. The first value of 5%, even unrealistic, was chosen to mimic the situation where the competing event recovery is rare. Other values chosen for the chance of recovery match the current incidence of recovery observed in real (clinical trials and cohort studies) research around the globe. Values reflecting how fast patients recover from COVID-19 expressed in terms of median time to recovery were chosen to vary between 5 to 20 days. Finally, the hazard parameter for death was chosen to be 0.05.

### Estimation of HR and 28-days ARR of in hospital mortality

Within each scenario and for each of the 1,000 generated samples, we estimated the HR and the 28-days ARR of in-hospital mortality using two models: the standard Cox PH and the FG competing risks model. The Cox PH model does not take recovery as a competing risk into account, while the FG model considers recovery as competing risk. The estimated quantities (HR and 28-days ARR) were then averaged across the 1,000 generated samples and compared to their corresponding true values. Patients who withdrew, who were lost to follow-up or who were still hospitalised at the end of the study were considered as right-censored in both Cox PH and FG models. All the simulations were carried out using the R software version 3.6.1^15^.

## 3. Results

### 3.1 Time to recovery and the bias of the estimated HR

For all the scenarios (except scenario 1) where the treatment has an effect on death, recovery, or on both, the estimated HR obtained from the Cox PH model were mis-estimated i.e. usually different from the true HR (Figure 1). For instance, in scenario 3 where the treatment has no effect on death but high effect on recovery, the HR estimated from the Cox PH model when the median time to recovery is around 10 days is 1.6 compared to 1 (the true HR) showing an over-estimation of 60% (Figure 1). Furthermore, the magnitude of this over-estimation increases with decreasing recovery time. This over-estimation disappears when the median time to recovery is very long, i.e. at the end of follow up when recovery is no longer a competing event. The results obtained from the FG model are, however, consistent in all scenarios and show no misestimation.

**Figure 1.**
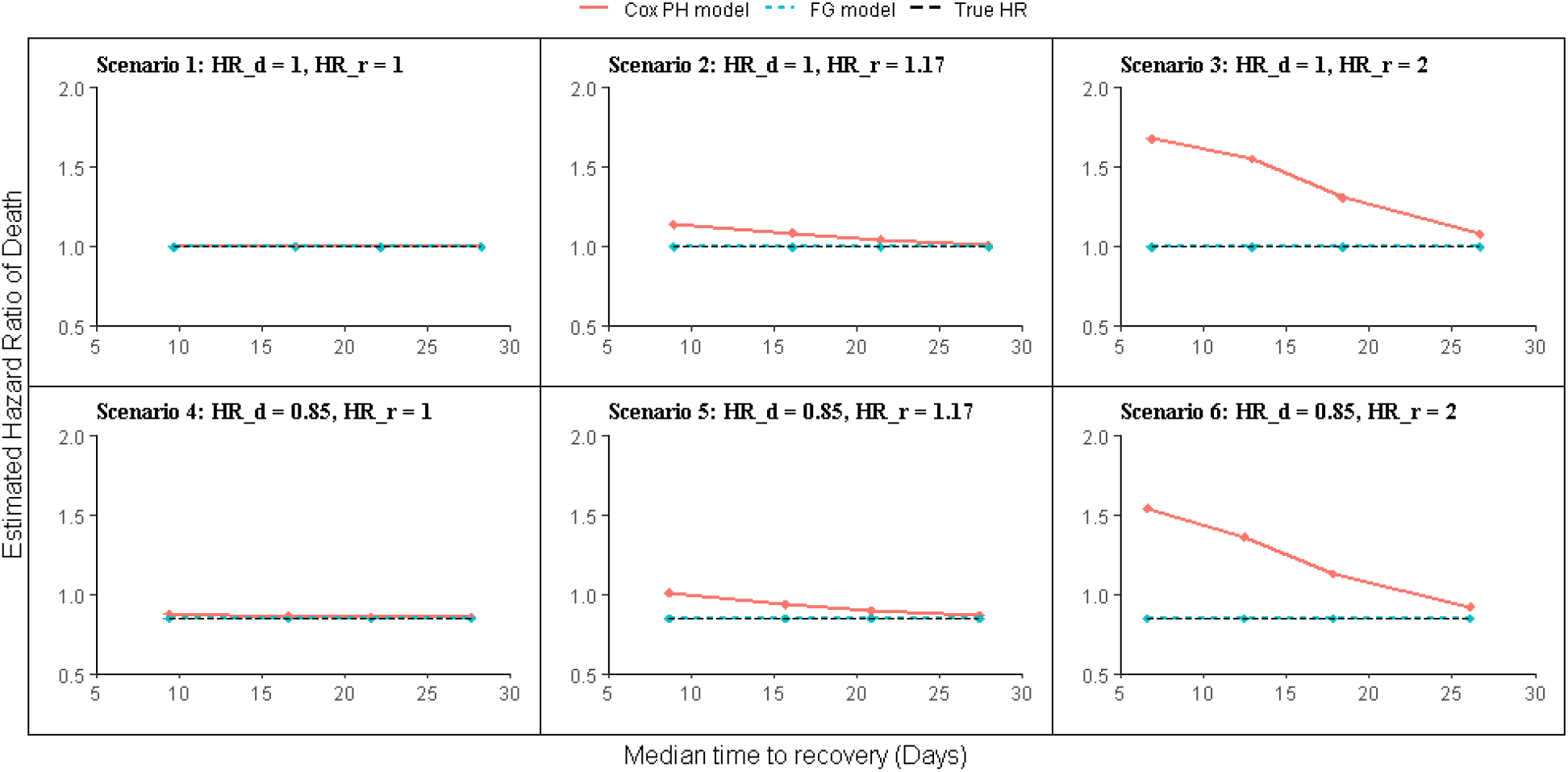
The HR estimated from the Cox PH model (red line) and the FG model (blue dashed line) across the six scenarios assuming a chance of recovery of 80%. Within each plot (scenario), the x-axis represents the median time to recovery where smaller values indicate faster time to recovery. The horizontal black dashed line within each plot represents the true HR.

### 3.2 Chance of recovery and the bias of the estimated HR

Figure 2 illustrates how the Cox PH model mis-estimates the true HR and how this mis-estimation increases in magnitude with increasing chance for recovery. For instance, in scenario 3 where the drug has no effect on death but high effect on recovery, and when the chance of recovery is around 90%, the HR estimated from the Cox PH model was around 2.1 compared to 1 (the true HR) showing a harmful effect of treatment with an over-estimation of 110% (Figure 2). More importantly, the estimated HR from the Cox PH model can not only over-estimating the true HR, but also incorrectly showing that the treatment is harmful when in fact it is beneficial. This is demonstrated in scenario 6 where treatment has a beneficial effect on both death and recovery but the effect on recovery is higher than the effect on death. In this scenario, the true HR for death is 0.85 indicating a beneficial effect of treatment (a reduction in the hazard of death of 15%). However, the estimated HRs from Cox PH model were 2.2, 1.83 and 1.55 when the chance of recovery were assumed to be 95%, 90% and 80%, respectively, showing incorrectly an increased risk of death in treated compared to placebo patients. The results obtained from the FG model are however consistent in all scenarios and show no mis-estimation or incorrect direction of the treatment effects (Figure 2).

**Figure 2.**
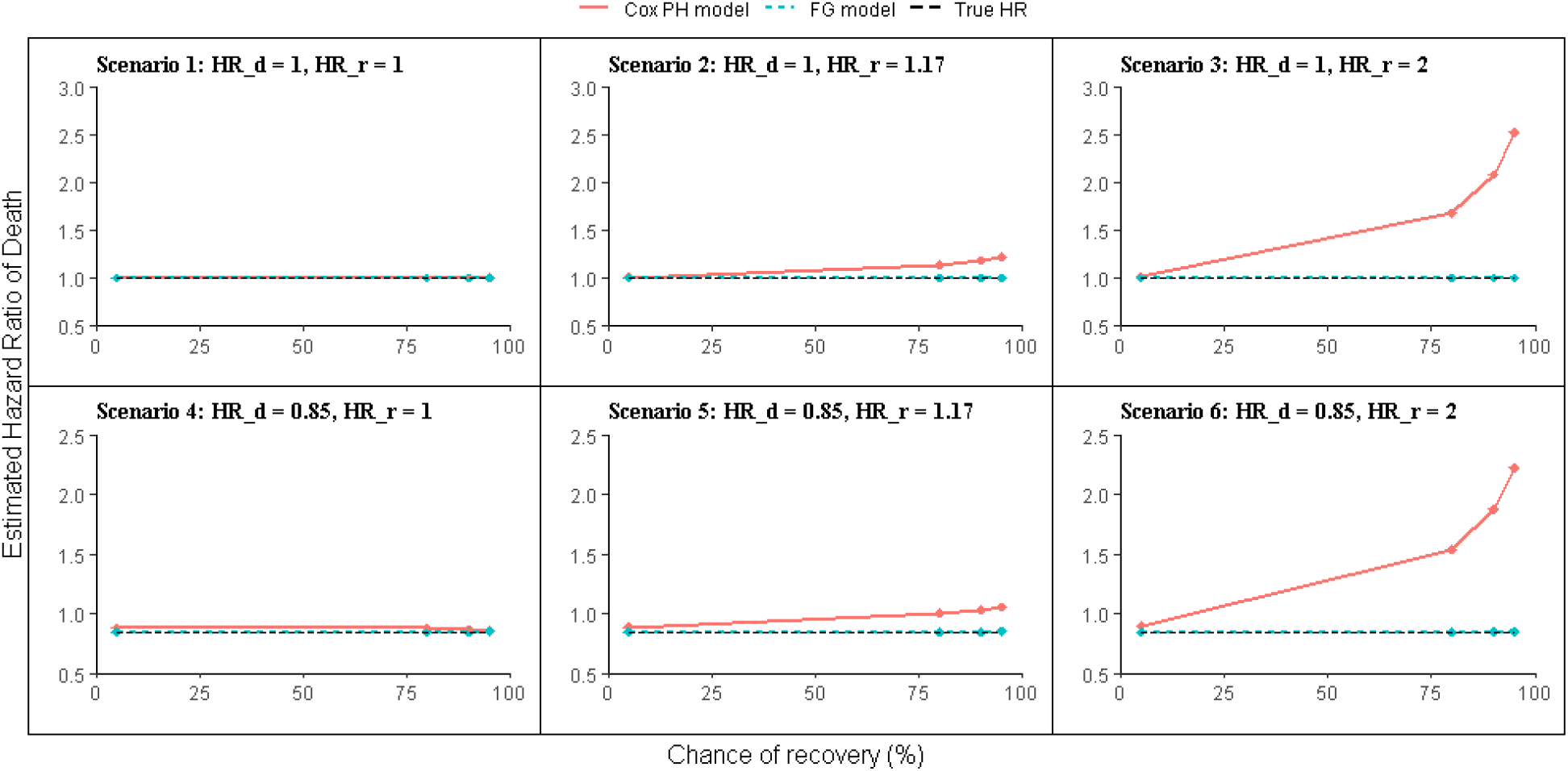
The HR estimated from the Cox PH model (red line) and the FG model (blue dashed line) across the six scenarios assuming a median time to recovery of around 10 days. Within each plot (scenario), the horizontal black dashed line represents the true HR and the x-axis represents the chance of recovery. The horizontal black dashed line within each plot represents the true HR.

### 3.3 Time to recovery and the estimated 28-day ARR of in-hospital mortality

Figure 3 shows how the Cox PH model misestimates the true 28-day ARR of in-hospital mortality and how this misestimation increases in magnitude when the median time to recovery decreases (i.e. when the process of recovery is faster). For instance, in scenario 3 where the drug has no effect on death but high effect on recovery, and when the median time to recovery is around 13 days, the 28-day ARR of in-hospital mortality estimated from the Cox PH model was around 12% compared to 0% (the true 28-day ARR). Interestingly, in scenario 6 where treatment has a beneficial effect on both death and recovery but the effect on recovery is higher than the effect on death, the Cox PH models not only over-estimated but also reversed the direction of effect of the 28-day ARR of in-hospital mortality. In this scenario, the true 28-day ARR for death is −2.5% showing a reduction in mortality of 2.5% in treated compared to placebo. However, the 28-day ARR estimated from the Cox PH model were +15%, +9% and +2.5% when the median time to recovery was 5 days, 13 days and 18 days, respectively, showing incorrectly an increased risk of death in treated compared to placebo patients. The results obtained from the FG model are however consistent across all scenarios and show no misestimation or incorrect direction of the treatment effects (Figure 3).

**Figure 3.**
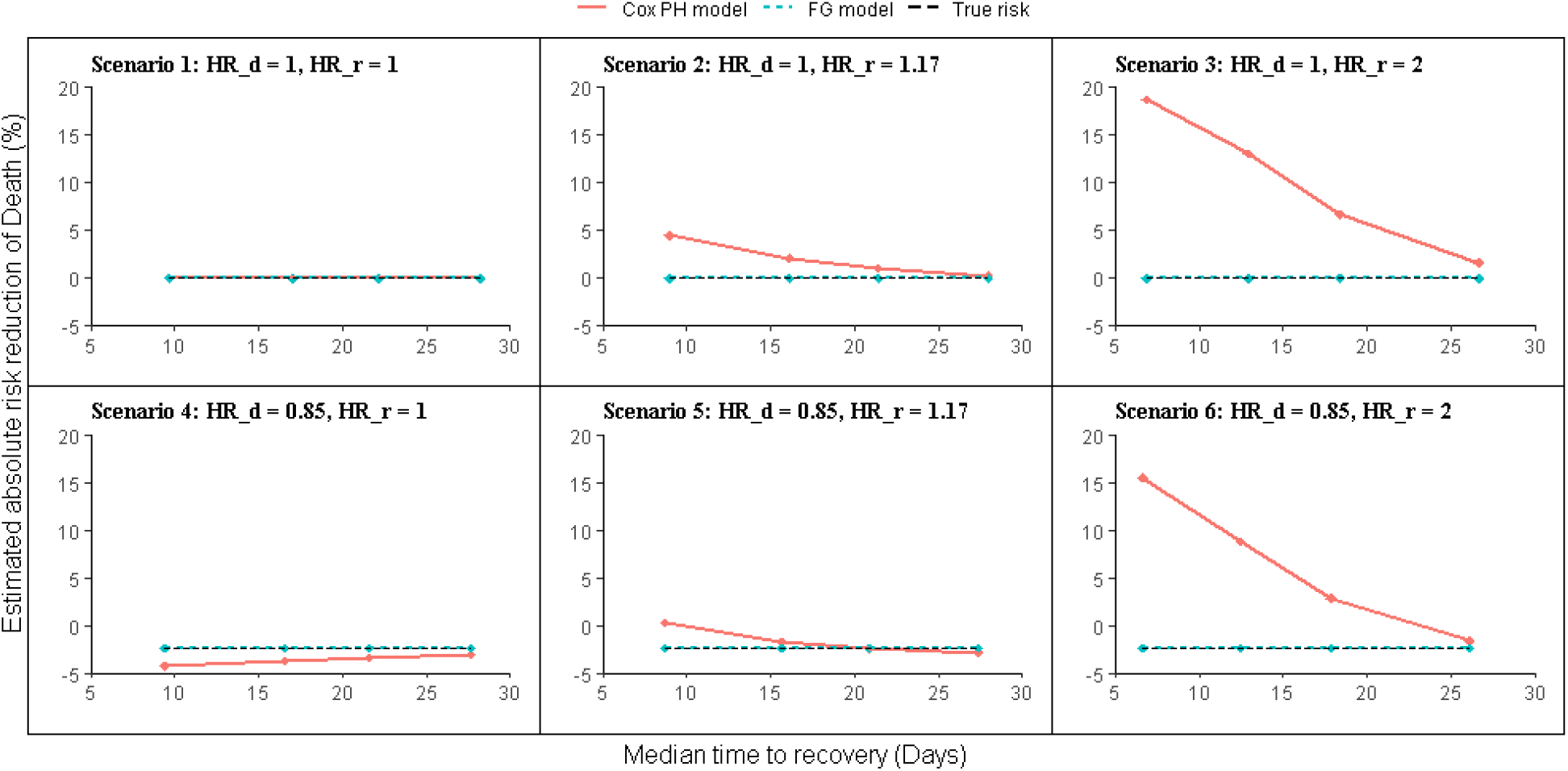
The 28-day ARR of in-hospital mortality estimated from the Cox PH model (red line) and the FG model (blue dashed line) across the six scenarios assuming a chance of recovery of 80%. Within each plot (scenario), the horizontal black dashed line represents the true 28-day ARR of in-hospital mortality and the x-axis represents the median time to recovery.

In scenario 1 of Figure 3, the 28-day ARR of in-hospital mortality estimated from the Cox PH model are similar to the true ones. This, however, does not exclude the possibility of a hidden mis-estimation of the 28-day risk of in-hospital mortality in both treated and placebo patients. As this misestimation is similar in both treated and placebo groups, the estimated 28-days ARR defined as the difference between the two quantities will be zero and therefore hides this misestimation. This is illustrated in figure 4 where the 28-day risk of death was estimated in both treated and placebo to 47% (when the median time to recovery was assumed to be 10 days) whereas the true risk is around 17% in both groups.

**Figure 4.**
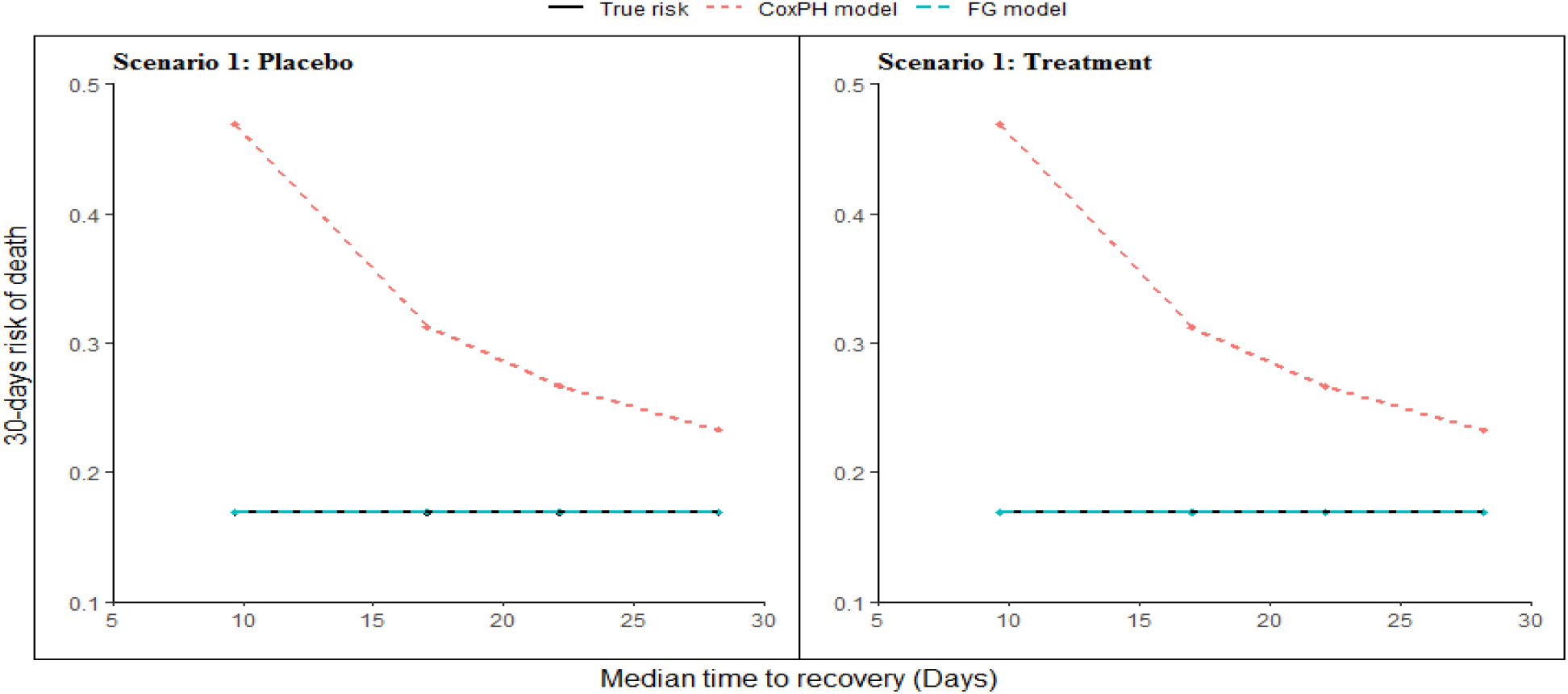
The estimated 28-day risk of in-hospital mortality in treated and placebo patients according to scenario 1 as a function of the median time to recovery. The red dashed line is for the Cox PH model, the blue line for the FG model and the black dashed line represents the true 28-day ARR.

## 4. Discussion

Using statistical modelling and extensive simulations mimicking real randomized clinical trials on COVID-19 similar to those involving the use of Hydroxychloroquine, Remdesivir and convalescent plasma therapy, this study discusses the impact of not taking the competing risk between death due to COVID-19 and recovery into consideration in survival analysis. The study provides clear evidence on how time to recovery and chance of recovery both affect the quality of the HR and the 28-day ARR of death estimated from the standard Cox PH model. It shows that ignoring the competing risks in survival analysis would affect the quality of the estimated HR and ARR leading to biased estimates. This effect is in particular pronounced if the chance of recovery is high and the time to recovery is short. This is of significant relevance, as in some cases the HR estimated by the standard Cox PH model could lead to the opposite direction as compared to the true HR. The study also shows that the FG model, that takes competing risks into consideration, performs largely better than the Cox PH model and provides estimates with no bias.

When in-hospital mortality is the primary outcome, considering patients who recovered at a given time point as right-censored, implicitly assumes that they have similar risk, of dying from COVID-19, compared to those who are still at risk (i.e. hospitalized) at that time point. Since a considerable proportion of COVID-19 patients recover, the concern of competing risk is inevitably of significant relevance. The same principle applies when recovery is the primary outcome.

Looking at currently published RCTs investigating potential treatment for COVID-19, most of the studies do use standard Cox PH models. Our simulations suggest that it would be helpful to confirm the findings of these trials with a competing risk analysis approach as the true effect of the studied drug, for example on death, may be misestimated or even reversed. In the recently published preliminary results from the RECOVERY trial for instance, the primary endpoint (28-day mortality) in the hydroxychloroquine arm was met in 25.7% and in the placebo arm in 23.5% with an estimated HR of death of 1.11^16^. Considering that the data on time to recovery are missing in this preliminary publication, the true HR for death may be misestimated based on our findings. It would therefore be of interest, to take competing risk into consideration for final analysis. Cao et al reported, in a trial investigating the effect of liponavir/ritonavir versus standard care in adults hospitalized for severe COVID-19, no benefit of the treatment in terms of the 28-day survival and time to clinical improvement^2^. The median time to discharge was 13 days and the statistical method chosen was the standard Cox PH model. Although there is no guarantee that using a competing risk analysis would have changed the results, the median time to discharge is within a time frame that could have impacted the neutral mortality effect observed in the trial based on our simulations.

Horby et al. randomised patients hospitalised for COVID-19 to either dexamethasone or usual care and report a reduction in the 28-day mortality, a benefit that was mainly observed among those receiving invasive mechanical ventilation or oxygen at randomization^17^. As the discharge rate from hospital within 28 days was 64.6% in the dexamethasone and 61.1% in the usual care group, respectively, again a rate that could have impacted the results.

Another candidate drug for the treatment of COVID-19 is Remdesivir, an inhibitor of the viral RNA-dependent, RNA polymerase in SARS-CoV2 and the Middle East respiratory syndrome (MERS-CoV)^18^. In a trial of 1,063 people comparing Remdesivir to placebo^19^ where mortality was analysed as a key secondary outcome, the reduction in the Remdesivir group did not reach statistical significance when the HR was derived from the Cox PH model (HR 0.70, 95%-CI 0.47 −1.04). In this setting, again a sensitivity analysis using a competing risk analysis approach would be more than welcome.

Although the simulation approach in our study was helpful to investigate the interplay of various hazards on death and recovery as well as time to recovery and to suggest a potential impact on currently ongoing COVID-19 research, we appreciate that we did not analyse actual trial data of patients with COVID-19. Hence, it would be critical to apply Cox PH and competing risk analysis approaches to data from already available randomised controlled trials.

Our study demonstrates that there is a substantial risk of misleading results in COVID-19 research if recovery and death due to COVID-19 are not considered as competing risk events. Therefore, we strongly suggest the use of competing risk approach in re-analysis of relevant published data to confirm the findings from standard Cox PH, and recommend the use of this approach in future COVID-19 research.

## Data Availability

This is a simulation study. No data on real patients was used. Upon justifiable request, the authors are happy to share the Data Generating Process (DGP) from which the simulations were carried out

## Acknowledgments

The authors would like to thank Prof. Geert Molenberghs for interesting discussions and suggestions that improved the paper.

